# Associations between depression symptom burden and delirium risk: a prospective cohort study

**DOI:** 10.1101/2023.09.21.23295926

**Authors:** Arlen Gaba, Peng Li, Xi Zheng, Chenlu Gao, Cai Ruixue, Kun Hu, Lei Gao

**Author notes:** Address correspondence to: Arlen Gaba, BS; Lei Gao, MBBS; Department of Anesthesia, Massachusetts General Hospital, Boston, MA, 02114.

## Abstract

**BACKGROUND AND OBJECTIVES:** Delirium and depression are increasingly common in aging. There is considerable clinical overlap, including shared symptoms and comorbid conditions, including Alzheimer’s disease (AD), functional decline, and mortality. Despite this, the long-term relationship between depression and delirium remains unclear. This study assessed the associations of depression symptom burden and its trajectory with delirium risk in a 12-year prospective study of older individuals during hospitalization.

**RESEARCH DESIGN AND METHODS:** 319,141 UK biobank participants between 2006-2010 (mean 58y [range 37-74, SD=8], 54% female) reported frequency (0-3) of four depressive symptoms (mood, disinterest, tenseness, or lethargy) in the preceding 2 weeks, and aggregated into a depressive symptom burden score (0-12). New-onset delirium was obtained from hospitalization records during 12y median follow-up. 40,451 (mean age 57±8; range 40-74y) had repeat assessment on average 8y after their first. Cox proportional hazard models examined whether depression symptom burden and trajectory predicted incident delirium during hospitalization.

**RESULTS:** 5,753 (15 per 1000) newly developed delirium during follow-up. Increased risk for delirium was seen for mild (aggregated scores 1-2, hazards ratio, HR=1.16, [95% confidence interval 1.08–1.25], *p*<0.001), modest (scores 3-5, 1.30 [1.19–1.43], *p*<0.001) and severe (scores ≥ 5, 1.38 [1.24–1.55], *p*<0.001) depressive symptoms, versus none in the fully adjusted model. These findings were independent of the number of hospitalizations and consistent across hospitalization settings (e.g., surgical, medical, or critical care) and specialty (e.g., neuropsychiatric, cardiorespiratory or other). Worsening depression symptoms (≥1 point increase), compared to no change/improved score, were associated with an additional 39% increased risk (1.39 [1.03–1.88], *p*=0.03) independent of baseline depression burden. The association was strongest in those over 65y at baseline (*p* for interaction <0.001).

**DISCUSSION AND IMPLICATIONS:** Depression symptom burden and worsening trajectory predicted delirium risk during hospitalization. Increased awareness of subclinical depression symptoms may be warranted for delirium prevention.

## Background and Objectives

Delirium is a cognitive insult characterized by its acute onset, fluctuating course of the reduction in attention and awareness, commonly occurring in hospital admissions as frequently as 50 percent of patients.^1,2^ Although delirium is a reversible form of cognitive impairment, it is associated with an increased risk for dementia, nursing home placement, functional decline, and mortality.^3^ Delirium has been linked to noncognitive features, such as sleep disruption^4^ and depression symptoms.^5^ There is a known overlap in symptoms and comorbid conditions between delirium and depression and a worse prognosis when both are present.^6,7^

In older hospitalized patients, depression symptoms can be present in up to half, depending on the population (medical vs. surgical) and measurement tools.^8,9^ Some evidence suggests that depression may be a risk factor for delirium.^10,11^ Yet, uncertainty remains regarding the long-term relationship between depression symptoms and delirium,^12,13^ particularly in larger population-based cohorts across therapeutic settings (e.g., general medical vs. postoperative) and age groups.^6^ In addition, shared comorbidities prevalent in older individuals, such as dementia or cardiometabolic disease, are also associated with delirium risk.^14,15^ Whether depression symptoms are a risk factor for delirium or a prodromal marker for neurodegeneration remains unclear.^16^

Given that depression symptoms are modifiable, our primary objective was to determine whether earlier life depression symptoms are a risk factor for incident delirium during hospitalization. Within a large community sample of middle- to older-aged adults from the UK Biobank, we examined the association between depression symptom burden derived from an aggregate symptom frequency score and new-onset delirium after hospitalization during a median 12 years of follow-up. We examined these relationships in clinically important subsets (postoperative delirium and after the exclusion of known dementia) and by common comorbidities. Finally, in a follow-up cohort, a median 4 years after the first assessment, we examined whether worsening depression symptom trajectory contributed to additional risk for delirium.

## Research Design and Methods

### Study participants and data resource

Between 2006 and 2010, over 500,000 aged 37 to 70 (57±8 years, 54% female) from across the United Kingdom were recruited to participate in the UK biobank. ^17^ Participants completed extensive questionnaires on demographics, lifestyle choices, medical conditions, and psychiatric well-being and were followed until February 2021 (median 12 years). 319,141 participants (mean age [SD]: 57.9 [7.9], range: 37.4-73.8 years; 54.0% female) completed psychological assessment with ≥1 hospitalization after baseline (given that delirium requires a precipitating illness event; Supplemental Fig. 1). A subset (n=40,451, 52% female, mean age 64±8; range 44-83y) was reassessed between 2012 and 2020 and followed for a median of 4 years. The UK Biobank structure and data validation efforts have been described in detail.^18^

### Standard Protocol Approvals, Registrations, and Patient Consents

The UK Biobank received National Research Ethics Approval, and participants gave written informed consent. This study was conducted under the terms of UK Biobank access number 40556 and Mass General Brigham IRB approval (#2020P002097).

### Screening of depression symptoms

Participants were asked about depression symptoms frequency with 4 questions; “Over the past two weeks, how often have you felt down, depressed, or hopeless?;” depressed mood), 2) “How often have you had little interest or pleasure in doing things?” unenthusiasm/disinterest), 3) “How often have you felt tense, fidgety, or restless?” tenseness/restlessness), and 4) “How often have you felt tired or had little energy?” tiredness/lethargy). We assigned scores to the responses: not at all (0), several days (1), more than half the days (2), or nearly every day (3).” A summed depression symptom score (0-12) was calculated for each participant, which we used to classify depression symptom burden in a way that keeps group power with increments of 2-points (representing one *significantly more* or two *slightly more* frequent symptoms) as follows: “none” (0), “mild” (1-2), “modest” (3-4) and “severe” (≥5). We excluded participants who responded with “do not know” or “prefer not to answer” [depressed mood (4.6%), unenthusiasm and disinterest (3.6%), tenseness and restlessness (4.2%), and tiredness and lethargy (3.1%)]. Depression symptom burden trajectory was calculated as the difference between the follow-up and baseline scores and categorized into “no change/improved” (≤0-point change) or “worsened” (≥1-point change). The distribution and change in scores are shown in Supplemental Figure 3.

### Assessment of delirium diagnosis

The UK biobank released linked hospitalization records and International Classification of Disease (ICD-10) diagnoses from the National Health Service during the follow-up period. Incident delirium was the first occurrence of the ICD-10 code F05, included in hospital admissions health records as described in previous studies.^4,19–22^ We excluded 61 cases where delirium predated the baseline assessment and 27 where delirium predated the follow-up assessment. The hospitalization settings of delirium, i.e., surgical (postoperative), medical (non-surgical), and critical care, were separately identified. We identified *postoperative delirium* (POD) using linked operation/procedure coding and matching operation dates within 3 days before delirium and tested in separate models.^23^ We classified a *medical* hospitalization setting as patients with delirium who did not have any associated operations or procedures. Finally, we identified those with delirium after admission to critical care units using critical care admission dates provided by the UK Biobank.

We further identified *non-dementia-related delirium* by excluding a subset of participants within the delirium group who had “delirium superimposed on dementia” (F05.1) or a prior diagnosis of any dementia. Admitting specialist/specialty was used to specify patients with delirium admitted to neuropsychiatric, cardiorespiratory, or other teams. Neuropsychiatric admitting specialty were found under the data field 41245, described as “Main Specialty of Consultant (recorded) Summary Administration.” See Supplemental Methods for specific grouping codes used.

### Assessment of covariates

Covariates were grouped based on 1) demographics, 2) lifestyle factors, 3) significant cardiovascular disease/risks (CVD)/comorbidities, and 4) Demographics, including age, sex, education, ethnic background, and controlling for number of hospitalizations post-assessment. Age at recent depression assessment was calculated in years based on the participants’ birth dates. Sex and ethnicity were self-reported at baseline. Ethnicity was included as European vs. non-European based on the distribution of participants of European descent (94%). Education was based on answering college attendance (yes/no).

Lifestyle factors included the Townsend Deprivation Index (TDI,) a material deprivation score and classified into higher/lower medians), physical activity (summed metabolic equivalent minutes), alcohol consumption (<4 drinks/≥4 drinks per week), BMI (weight [kg] divided by the height squared [m^2^]), sleep duration was categorized into short (<6h/day), normal (6-9h), and long (>9h) because of the previously demonstrated U-shape associations with delirium or dementia,^4,24^ frequency of friend and family visits (never vs. any), and falls in the last year (none vs. any).

CVD was based on hypertension, high cholesterol, smoking, diabetes, ischemic heart disease, and peripheral vascular disease. Comorbidities included a previously described morbidity burden^21,25,26^ based on the summed presence of any cancers, respiratory, neurological, gastrointestinal, renal, hematological, endocrine, musculoskeletal, connective tissue, infectious diseases/disorders, and classified as none (0)/modest (1-3)/high (≥4) conditions. Cognitive performance was estimated at initial enrollment using a raw processing speed test involving the mean reaction time to identify card matches correctly.^27^

The full final model included serum 25-hydroxyvitamin D (25[OH]D, a proxy for vitamin D levels recently linked to delirium within this cohort, categorized into sufficient >50nmol/L, low 25-50nmol/L, and deficient <25nmol/L), and pre-existing dementia/Parkinson’s disease, or depression diagnosis/treatment (any from seeing a psychiatrist, use of antidepressants, or a self-reported/ICD-10 diagnosis).

### Statistical Analysis

The features of those who developed delirium compared to those who were hospitalized but remained delirium-free during follow-up were compared using chi-squared tests for categorical variables (e.g., sex, ethnicity, presence/absence of comorbidities, recent smoking) and independent samples t-tests or the non-parametric, Kruskal–Wallis for continuous variables (e.g., age, BMI, TDI, physical activity, reaction time, CVD, depressive symptoms burden score, frequency of falls in the last month). Cox proportional hazard models were used to evaluate the association between depressive symptoms burden and time to incident delirium [reported as hazard ratios (HRs) and corresponding to 95% confidence intervals (CIs)].

The core model (A) controlled for demographics (age, sex, college education, ethnicity, and number of hospitalizations). The lifestyle model (B) additionally controlled for TDI, physical activity, alcohol consumption, BMI, sleep duration, frequency of family and friend visits, and falls in the last year. The significant CVD/Comorbidities model (C) further controlled for CVD risk score, morbidity burden, and cognition. The final model (D) controlled for vitamin D levels, Parkinson’s/Dementia, and depression diagnosis. We again examined the association between depression symptom burden in the follow-up cohort. Using the core model, we adjusted for the baseline depressive score and the time-lag between assessments. Sensitivity analysis examined the relationship between the depression score and post-operative (surgical), medical (non-surgical), non-dementia-related delirium, critical care delirium in the full cohort in addition to admitting specialty in delirium cases separated into neuropsychiatric, cardiorespiratory and others (non-cardiorespiratory, non-neuropsychiatric related admissions). Time-to-event was the years between depressive symptoms assessment and delirium diagnosis. Delirium-free participants were censored in February 2021, the last date of available records. All other statistical analyses were performed using JMP Pro (Ver. 16, SAS Institute, Cary, NC, USA). *P* value < 0.05 was used for statistical significance.

Data are available from the UK Biobank after submitting an application. The syntax for conducting the analysis is available upon reasonable request.

## Results

### Participant characteristics

This prospective study included 319,141 participants mean [SD] age: 57.9 [8.0], range: 37.4-73.8 years; 54.0% female) who had all data available, were hospitalized at least once after the first assessment, and had no prior delirium. (Supplemental Fig. 1). The cohort was followed for a median period of 12.0 years (IQR 11.2–12.7) after baseline depression symptom burden assessment. Within this period, 5,753 (15 per 1000) developed delirium.

Participants with incident delirium were more likely to be older (64.0 years vs. 57.9 years), male (57.3% vs. 45.7%), lower chance of college attendance 20.8% vs. 30.0%), were more likely to be of European ancestry (95.4% vs. 94.1%), lived in areas of greater deprivation (TDI -0.62 vs. - 1.30), had higher BMI (28.7 vs. 27.7) than those who remained delirium-free. The incident delirium participants were less active (1962.5 vs. 2079.4 met-minutes), did not have differences in alcohol consumption, and were more likely to sleep outside the recommended 6-9h range (<6 h/day: 8.2% vs. 6.1% and >9h/day: 4.5% vs. 2.1%), had a higher percentage of no family visits that year (3.4% vs. 1.8%), and more likely to have fallen that year (31.6% vs. 21.4%). The delirium group was more likely to have one or more CVD (68.9% vs. 31.1%), higher morbidity burden with 4 or more conditions (40.5% vs. 32.0%), higher incidence of dementia/Parkinson’s disease (2.5 % vs. 0.2%), slower reaction time (613ms vs. 559ms), and more likely to be vitamin D deficient (5.7% vs. 3.7%). Participants with delirium were also diagnosed or self-reported depression more (10% vs. 7%) and had more of the cohort in the severe category of depressive symptom burden scoring (12.2% vs. 9.4%) (Table 1).

**Table 1.** Demographics, lifestyle, and clinical comorbidities at baseline.

|  | Developed new-onset<br>delirium<br>(n = 5,753)<br>Mean (SD), or n (%) | Did not develop<br>delirium<br>(n = 319,141)<br>Mean (SD), or n (%) | *p value* |
| --- | --- | --- | --- |
| **Demographics** |  |  |  |
| Age at baseline | 64.0 (5.4) | 57.9 (7.9) | <0.001 |
| Males | 57.3% | 45.7% | <0.001 |
| College Attendance | 20.8% | 30.0% | <0.001 |
| Ethnic Background (European) | 95.4% | 94.1% | 0.001 |
| Townsend deprivation index <sup>a</sup> | -0.62 (0.04) | -1.30 (0.01) | <0.001 |
| Body mass index (kg/m<sup>2</sup>) | 28.7 (5.4) | 27.7 (4.9) | <0.001 |
| **Lifestyle** |  |  |  |
| Physical activity (MET-mins) <sup>b</sup> | 1963 (493) | 2079.4 (462) | <0.001 |
| Alcohol (≥ 4 drinks/week) | 47.5% | 47.0% | 0.43 |
| Sleep Duration |  |  |  |
| short (<6h/day) | 8.2% | 6.1% | <0.001 |
| normal (6-9h) | 87.4% | 91.9% | <0.001 |
| long (>9h) | 4.5% | 2.1% | <0.001 |
| Frequency of family visits (never) | 3.4% | 1.8% | <0.001 |
| Falls in the last year (any) | 31.6% | 21.1% | <0.001 |
| **Comorbidities** |  |  |  |
| CVD risk <sup>c</sup> (any) | 68.9% | 31.1% | <0.001 |
| Morbidity Burden (high) | 40.5% | 32.0% | <0.001 |
| Dementia/Parkinson's disease | 2.5% | 0.2% | <0.001 |
| Cognition (reaction time) <sup>d</sup> | 613 (145) | 559 (118) | <0.001 |
| Vitamin D (deficient) <sup>e</sup> | 5.7% | 3.7% | <0.001 |
| **Depression/anxiety** <sup>f</sup> | 10.0% | 7.0% | <0.001 |

Note. Participant characteristics at baseline by delirium status. <sup>a</sup>higher value = worse deprivation. <sup>b</sup>METS-min/week increase. <sup>c</sup>CVD risk score: summed hypertension, cholesterol, diabetes mellitus, smoking status, and ischemic heart disease. <sup>d</sup>Cognition reaction time in milliseconds: average timed tests of symbol matching. <sup>e</sup>Vitamin D levels: sufficient >50nmol/L, low 25-50nmol/L, and deficient <25nmol/L). <sup>f</sup>Participants self-reported depression and anxiety symptoms, or ICD depression diagnosis. *SD* standard deviation, *CVD* cardiovascular disease.

### Depressive symptoms and associations with incidence of delirium

Figure 1A shows a stepwise increase in risk for the first occurrence of delirium with increasing depression symptom burden (mild, modest, and severe vs. none) for the core model. This translated into a higher cumulative incidence of delirium over the follow-up period (Figure 1B). Compared to no depressive symptoms, those with mild (HR=1.16, 95% CI [1.08-1.25], *p*<0.001), modest (1.30 [1.19-1.43], *p*<0.001), or severe (1.38 [1.24-1.55], *p*<0.001) depressive burden remained at higher risk for delirium in the fully adjusted model (Table 2). Using coefficients (ratio of the natural log of HRs) from the core model (Supplemental Table 1), the risks of modest and severe depression burden were equivalent to the effects of an additional 4 and 7 years of aging, respectively.

**Figure 1.**
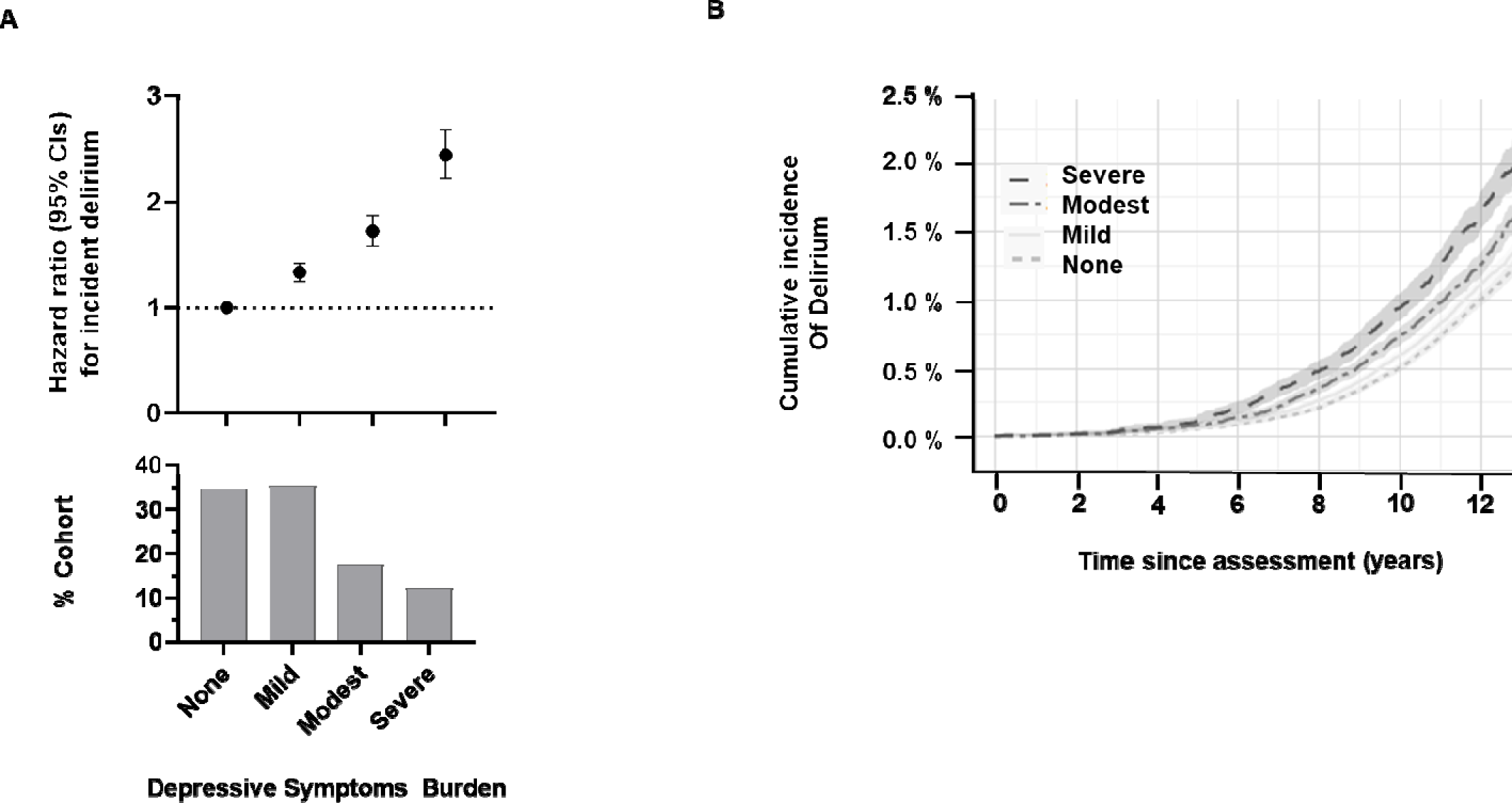
Depressive symptoms burden groups and risk for incident delirium. **(A)** Unadjusted cumulative incidence plot showing the percentage of the cohort with a first diagnosis of delirium over time, in the four depressive symptoms categories (None =0, Mild =1-2, Modest=3-4, and Severe risk =≥5), based on the depression symptom burden score. Hazard ratios (±95% CI) for incident delirium using Cox proportional hazards regression models adjusted for age, sex, education, and ethnicity, percentage of the cohort by depression symptom burden group in the panel below. **(B)** Cumulative incidence plot showing the percentage of the cohort with a first diagnosis of delirium over time in the four depressive symptom burden groups.

**Table 2:** Depressive symptoms burden and associations with incident delirium.

|  | All Delirium<br>N = 5753 | Postoperative Delirium<br>N = 1689 | Non-Dementia Related<br>N= 4064 |
| --- | --- | --- | --- |
|  | HR (95% CI)<br>*p*-value | HR (95% CI)<br>*p*-value | HR (95% CI)<br>*p*-value |
| Model A |  |  |  |
| Mild | 1.30 (1.22 – 1.38)<br><0.001 | 1.29 (1.15 – 1.45)<br><0.001 | 1.34 (1.24 – 1.44)<br><0.001 |
| Modest | 1.70 (1.58 – 1.84)<br><0.001 | 1.71 (1.48 – 1.96)<br><0.001 | 1.75 (1.60 – 1.92)<br><0.001 |
| Severe | 2.43 (2.22 – 2.68)<br><0.001 | 2.26 (1.92 – 2.66)<br><0.001 | 2.58 (2.33 – 2.86)<br><0.001 |
| Model B |  |  |  |
| Mild | 1.24 (1.16 – 1.32)<br><0.001 | 1.22 (1.09 – 1.37)<br>0.0007 | 1.23 (1.15 – 1.33)<br><0.001 |
| Modest | 1.44 (1.32 – 1.57)<br><0.001 | 1.50 (1.30 – 1.73)<br><0.001 | 1.45 (1.32 – 1.60)<br><0.001 |
| Severe | 1.70 (1.53 – 1.88)<br><0.001 | 1.67 (1.40 – 2.00)<br><0.001 | 1.75 (1.56 – 1.96)<br><0.001 |
| Model C |  |  |  |
| Mild | 1.19 (1.11 – 1.27)<br><0.001 | 1.17 (1.04 – 1.32)<br>0.008 | 1.18 (1.09 – 1.27)<br><0.001 |
| Modest | 1.35 (1.24 – 1.47)<br><0.001 | 1.40 (1.21 – 1.63)<br><0.001 | 1.36 (1.24 – 1.49)<br><0.001 |
| Severe | 1.48 (1.33 – 1.64)<br><0.001 | 1.45 (1.21 – 1.75)<br><0.001 | 1.50 (1.34 – 1.69)<br><0.001 |
| Model D |  |  |  |
| Mild | 1.16 (1.08 – 1.25)<br><0.001 | 1.16 (1.02 – 1.30)<br>0.02 | 1.15 (1.07 – 1.25)<br><0.001 |
| Modest | 1.30 (1.19 – 1.43)<br><0.001 | 1.39 (1.19 – 1.63)<br><0.001 | 1.30 (1.18 – 1.44)<br><0.001 |
| Severe | 1.38 (1.24 – 1.55)<br><0.001 | 1.44 (1.18 – 1.75)<br><0.001 | 1.44 (1.28 – 1.62)<br><0.001 |

Note. Cox proportional hazard models examining the association between depressive symptoms groups (with None as reference) and all delirium cases, and subgroups. Model A: demographics. Model B additionally includes Townsend deprivation index, physical activity, alcohol consumption, body mass index, sleep duration, frequency of friend and family visits, and falls. Model C: cardiovascular risk, morbidity burden, and reaction time. Model D: dementia/Parkinson's, vitamin D, and depression/anxiety diagnosis. *HR* hazard ratio, *CI* confidence intervals.

These results remained consistent when considering postoperative delirium only, after excluding known dementia (i.e., non-dementia-related diagnoses of delirium), medical (non-surgical), and critical care. Furthermore, these results were consistent in all cardiorespiratory admitting teams, neuropsychiatric, and all other non-neuropsychiatric or cardiorespiratory admitting teams (Supplemental Figure 2.) The effects of individual depression symptoms (daily vs. none) are presented in Supplemental Table 2. Those with depressed mood (2.17 [1.84–2.55, *p*<0.001), unenthusiasm/disinterest (1.88 [1.61–2.21], *p* <0.001), tenseness/restlessness (2.25 [1.84-2.63], *p* <0.001), tiredness/lethargy (2.48, [2.26–2.72], *p* <0.001) were all at increased risk for incident delirium. However, greater attenuation was seen for those reporting “depressed mood” and the anhedonia-like question on “unenthusiasm/disinterest” in the final models.

### Depression symptoms trajectory and risk for delirium

In the follow-up cohort of 40,451 participants, 213 (5.3 per 1000) developed incident delirium (median follow-up time: 3.8 years [range 11 months to 11.2 years; SD 2.7]). The median time from the initial depressive symptoms screening was 8.0 years [range 2.6-13.8 years; SD 2.7 years]. After adjusting for demographics, those who reported mild (1.51 [1.12-2.05], *p*=0.008), modest (1.74 [1.13–2.67], *p*=0.01), and severe (2.80 [1.63–4.80], *p*<0.001) depression symptoms were again associated with increased delirium risk when compared to those reported none (Table 3). After adjusting for participant baseline depression symptoms burden score and time-lag, a worsening score (≥1) depression symptoms burden score was associated with an additional 39% increased risk (1.39 [1.03–1.88], *p*=0.03; Table 3) compared to those reporting no change/improved score. To mitigate the ceiling effect (those scoring high at baseline have no room for worsening), we tested only those within the none and mild groups (baseline depression score 0-2, 74% of the cohort) and confirmed that a worsening score (≥1), was associated with an increased risk (1.45 [1.04–2.02], *p* = 0.03, Supplemental Table 3).

**Table 3.**
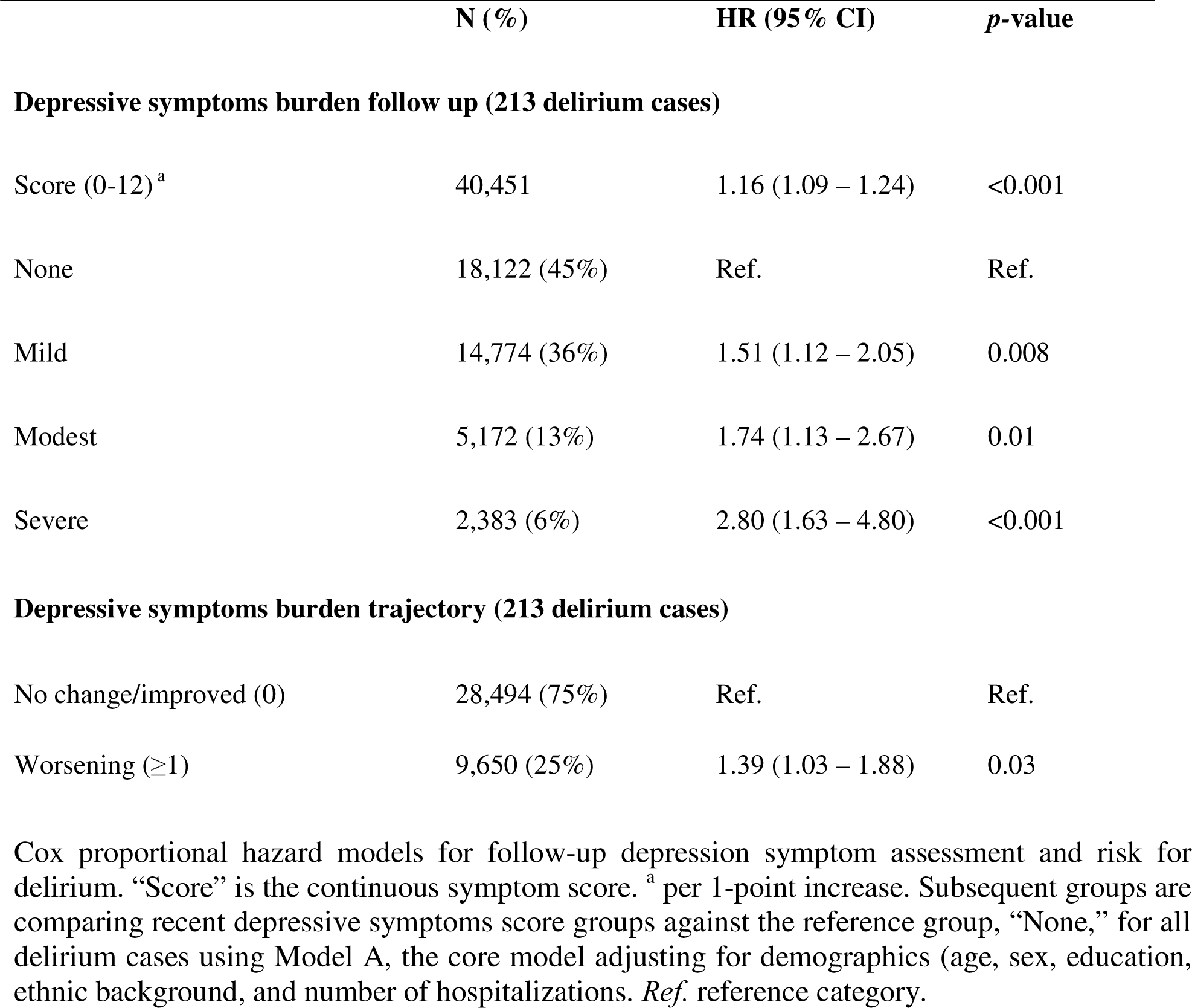
Demographics, lifestyle, and clinical comorbidities at baseline.

|  | N (%) | HR (95% CI) | p-value |
| --- | --- | --- | --- |
| **Depressive symptoms burden follow up (213 delirium cases)** |  |  |  |
| Score (0-12) <sup>a</sup> | 40,451 | 1.16 (1.09 – 1.24) | <0.001 |
| None | 18,122 (45%) | Ref. | Ref. |
| Mild | 14,774 (36%) | 1.51 (1.12 – 2.05) | 0.008 |
| Modest | 5,172 (13%) | 1.74 (1.13 – 2.67) | 0.01 |
| Severe | 2,383 (6%) | 2.80 (1.63 – 4.80) | <0.001 |
| **Depressive symptoms burden trajectory (213 delirium cases)** |  |  |  |
| No change/improved (0) | 28,494 (75%) | Ref. | Ref. |
| Worsening (≥1) | 9,650 (25%) | 1.39 (1.03 – 1.88) | 0.03 |

Cox proportional hazard models for follow-up depression symptom assessment and risk for delirium. “Score” is the continuous symptom score. <sup>a</sup> per 1-point increase. Subsequent groups are comparing recent depressive symptoms score groups against the reference group, “None,” for all delirium cases using Model A, the core model adjusting for demographics (age, sex, education, ethnic background, and number of hospitalizations. *Ref.* reference category.

### Incident delirium risk by subgroups

The risk of delirium was further examined by age (<65y/≥65y), sex, physical activity (lower/higher), morbidity burden, depression, reaction time, and sleep duration (Figure 2). Comparing participants with modest/severe vs. no depressive symptoms, those aged ≥65y were more strongly associated with delirium risk (1.70 [1.56-1.86]) compared to participants aged <65y (1.36 [1.24-1.48]) *p* for interaction <0.001. Similarly, patients without a depression diagnosis were more strongly associated (1.65 [1.54-1.76]) compared to those with diagnosed depression (1.29 [1.07-1.54]), *p* for interaction <0.001. Depression symptom burden was equally predictive in males and females, those with above average and below average physical activity, morbidity risk, reaction times, and nighttime sleep duration.

**Figure 2.**
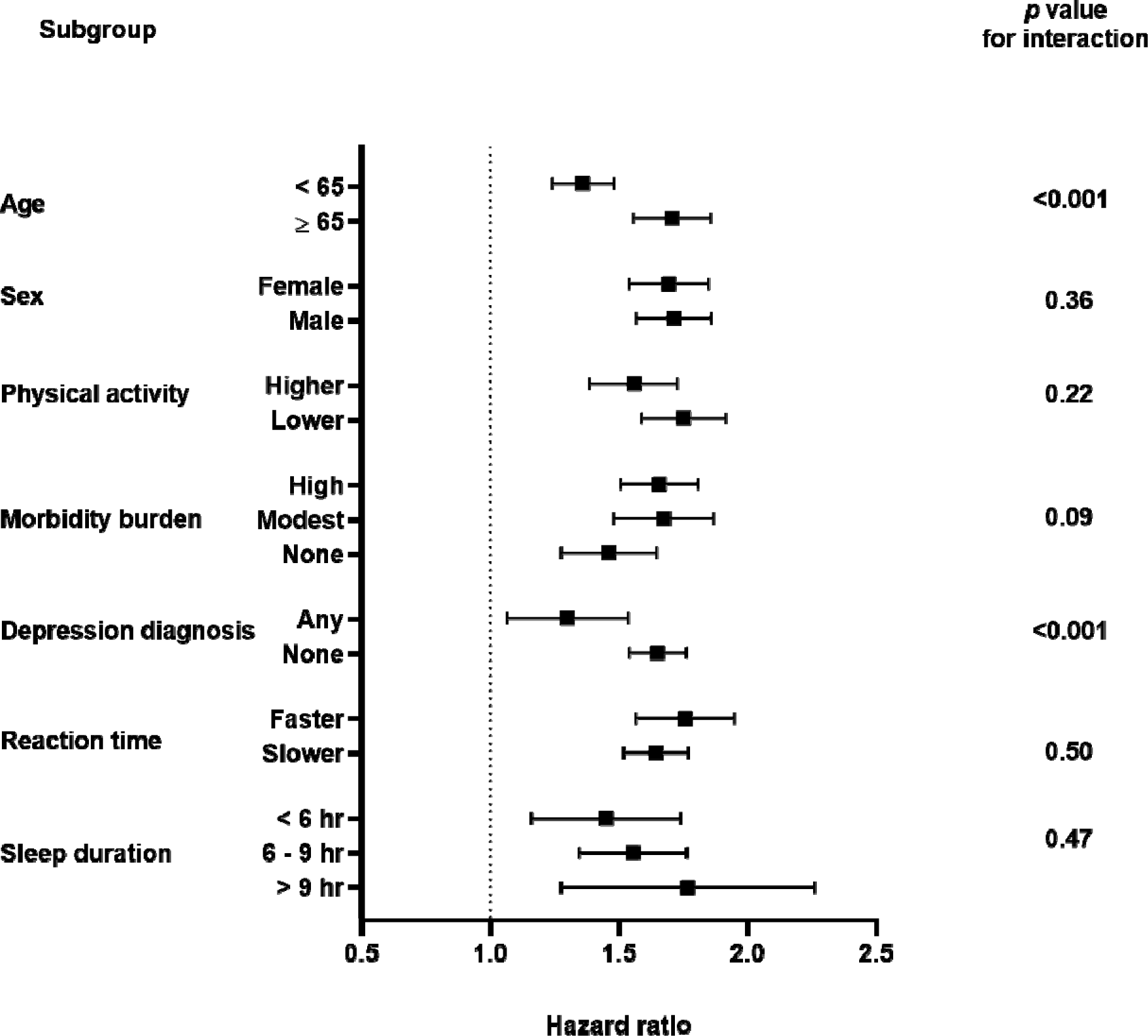
Forrest plot of hazard ratios and 95% confidence intervals for modest/severe depression symptoms burden (versus none/mild) predicting incident delirium based on subgroups of participants by age, sex, physical activity, morbidity burden, diagnosed depression, reaction time and night-time sleep duration.

## Discussion and Implications

Our study of 319,141 community-based UK Biobank participants found that those reporting mild, modest, and severe depression symptom burden were at 16%, 30%, and 38% higher risk for developing hospital-diagnosed delirium over a median 12 years of follow-up when compared to those reporting none. The findings were consistent for postoperative delirium and after the exclusion of underlying dementia, our main secondary analysis. In non-postoperative and critical care settings, these results remained consistent. Further sensitivity analysis of consulting/admitting specialty demonstrated that depressive symptoms were equally associated with incident delirium in neuropsychiatric, cardiorespiratory, or other admissions. More recent reporting in a smaller follow-up cohort of 40,451 confirmed the association between depression symptoms and delirium risk; in fact, those reporting worsened depression symptom trajectory were at an additional 39% risk. The association was strongest when depression symptoms were reported after the age of 65 years and in individuals without a history of depression/anxiety.

These findings are consistent with prior work showing that psychiatric well-being measures reliably predict delirium development.^11,28^ Specifically, postoperative delirium is more likely in those with baseline depression and depressive symptoms.^10,29,30^ Dysphoric mood and hopelessness, as components of depression symptoms, also increased the risk for delirium.^28^ While all components/questions drove these results, interestingly, tenseness/restlessness and tiredness/lethargy were most strongly associated with delirium in the final models, suggesting that the full spectrum of depression and anxiety-related symptoms reported should be considered.^9^ The simplicity of the assessment questions captured responses on a large scale, allowing for repeated measures and examining symptom trajectory. Consistent results across two separate time points and the additional risk from worsening symptoms support the idea that depression may increase neurocognitive vulnerability to stressors such as illness, surgery, or hospitalization rather than simply being comorbid with delirium. If replicated, these findings suggest the need for optimizing depression symptom burden in older adults, separately or as part of established multicomponent delirium bundles. For example, despite consistency in our findings across hospitalization settings (Supplemental Figure 2), there is a window of opportunity before major surgery to intervene,^9^ given that delirium is growing in an aging population with exponential increases in surgical needs.^31^

Whether these results point to a causal role or an unmasking of cognitive vulnerability is unclear. Underlying diseases linked to depressive symptomatology may contribute, despite being included in our models. Dementia is commonly comorbid with both delirium and depression.^6,14^ Neurophysiological disturbances in delirium include aberrations in monoamine neurotransmission and the imbalance of dopaminergic and cholinergic signaling.^32^ Twin studies have found an association between serotonin 2A receptor gene promoter A/A genotype and depression in older men.^33^ While late-life depression is associated with Alzheimer’s dementia, causal links have not been established.^34^ Other mechanisms include shared vulnerability to inflammation after illness or surgery and the impact on the aging brain and the endocrine system. Elevated endogenous cortisol levels have been observed for depression and in patients with severe dementia and delirium,^35,36^ and implicated in delirium pathophysiology.^37,38^ Finally, depression and dementia are often accompanied by sleep and circadian disruptions.^39,40^ In this study, differences in sleep duration did not modify the association between depression symptoms and delirium. Fluctuations of symptoms and intensity of delirium suggest an altered circadian rhythm.^41^ Recent evidence also indicates that circadian disturbances predispose to delirium,^25^ suggesting a bidirectional relationship. In this study, we accounted for sleep duration, and the association between depression symptom burden and delirium remained after controlling for known dementia and excluding preexisting dementia (or “delirium superimposed on dementia” cases). However, the interplay between depression, sleep/circadian health, and delirium risk in the older population is an emerging area of interest.^42,43^

The association between delirium and depression symptom burden (significant vs. mild/none) was strongest in older participants over 65y (vs. <65y) and in those *without* a depression diagnosis (vs. those with). Although age is one of the strongest independent risk factors for developing delirium, this suggests that concurrent depression burden is even more important to identify in older persons when preventing delirium. One interpretation is that cognitive impairment was underreported in the older cohort and not adequately controlled. Another possibility is that the temporal burden of depression symptoms, which may have been undertreated or underrecognized in those over 65, was not accounted for. This could also apply to participants without a formal diagnosis of depression but still reported significant symptoms, which may have been left untreated, leading to greater delirium vulnerability. Our findings of an increased risk in those with a worsening trajectory of depression symptoms support the latter. Unfortunately, details on treatments were not available in this study. Screening questions in this study may be more sensitive in detecting symptoms in patricians without a diagnosis. While caution is needed, these results emphasize the importance of addressing older adults’ psychiatric well-being, even in the absence of depression/anxiety diagnosis, to enhance neurocognitive reserve in response to acute illness or major surgical procedures. Depressive symptoms should not be regarded as a normal response to aging, as they have neurocognitive consequences.^44^

Strengths of this study include large sample size, long prospective follow-up, and repeat assessment. The sample sizes dedicated to delirium are also uniquely large.^45^ However, there are several limitations. UK Biobank participants are mostly Caucasian of European descent and may have healthier behaviors than the general UK population. This may underestimate the associations since participants agreeing to participate may have healthier habits, fewer comorbidities, and lower rates of psychiatric burden and delirium. For example, the interpretation of dysphoria and other aspects of psychiatric well-being may vary across different ethnicities and socioeconomic backgrounds, cautioning against extrapolating these findings to populations outside this specific demographic. Despite this, prior work has shown that risk factor associations in the UK Biobank are generalizable.^46^

The questionnaire items were selected by a UK Biobank working group consensus of experts that needed to balance broad utility with low patient burden given the large sample size.^17^ This study employed a brief rating scale using four items related to the patient health questionnaire to asses psychiatric well-being.^6,29,47–49^ The simplicity allows faster assessment on a large scale, but it is not a complete evaluation. The repeat assessment for depression symptom trajectory is limited in power and subject to selection bias in those who agreed to be reassessed. Furthermore, follow-up trajectories may be affected by ceiling and floor effects (e.g., quantifying changes in individuals with none or maximum depression symptom burden at baseline is not possible with our fixed scale).

We controlled for a wide range of confounders and stratified by subgroups. Still, there is likely residual confounding in the described relationships, given the complex nature of depression symptomatology and heterogeneity of delirium. Although we were able to adjust our models for one cognitive test, UK Biobank does not have other cognitive measurements, such as the Mini-Mental State Examination. We cannot exclude the possibility that many with delirium had undiagnosed cognitive impairment that we could not adjust for. Those with subclinical depression and depressive symptoms may have had maladaptive behaviors and consequences (e.g., poor stress tolerance, coping strategies, higher chance of future substance use, lack of social support), increasing opportunities for delirium via increased hospitalization numbers or presenting diagnoses more likely to precipitate delirium even when controlling for number of hospitalizations during follow-up. Given the potential relationship of those confounding factors with our exposure (i.e., depression), we grouped participants by admitting specialty physician/primary team as a proxy for admitting diagnosis (neuropsychiatric and cardiorespiratory) and hospitalization setting (post-operative, non-operative, critical).

Carefully designed longitudinal studies tracking depression/anxiety symptoms before hospitalization, e.g., a planned, elective major surgery, would help to confirm our observed link between depression symptoms and delirium. On the other hand, our multivariable-adjusted models may have accounted for covariates that could be on the causal pathway, e.g., physical activity and alcohol/substance use. Changes in these factors, driven by depression symptoms, can potentially impact delirium risk. Therefore, the results may underestimate the true strength of the relationship. Finally, clinical data in the UK Biobank cohort was limited to ICD coding. Others have used this approach for delirium,^50^ within this cohort and are highly specific (up to 96%) for delirium,^51^ but the sensitivity is modest (53-64% in recent studies).^51,52^ Thus, we are likely missing cases, particularly milder or hypoactive forms.

Our findings provide evidence bridging psychiatric well-being and delirium prevention. Since depression symptoms are modifiable and a non-cognitive proxy for resilience to inciting stressors before delirium, it may prove useful for neurological risk stratification alongside traditional risk factors. Additional work is required to determine the underlying mechanisms and whether a causal relationship exists before focusing on screening and treatment.

## Acknowledgments

This research has been conducted using the UK Biobank Resource under Application Number 40556.

## Author Contributions

Conception and design of the study: Gaba, Li, Hu and Gao L Acquisition and analysis of data: Gaba, Li, Zheng, Hu, and Gao L. Drafting a significant portion of the manuscript or figures: Gaba, Li, Gao C, Cai R, Hu, and Gao L.

## Supplemental Methods

Included specialty of the consultant in our neuropsychiatric cohort were coded such as 1020 Adult mental illness, 1270 Forensic Psychiatry, 1150 Clinical neuro-physiology, 1500 Neurology, and other specialties that fit the criteria, capturing diagnoses such strokes, TIA and acute psychiatric illness. Cardiorespiratory admitting specialty codes were specialties such as 1070 cardiology, 1080 cardiothoracic surgery, 1800 respiratory medicine and others that fit the criteria, capturing diagnoses such as heart failure, ischemic heart disease, pneumonia and others. “Others,” were all other specialty encodings that did not fit into these two specifications.

## Supplemental Tables

**Supplemental Table 1.**
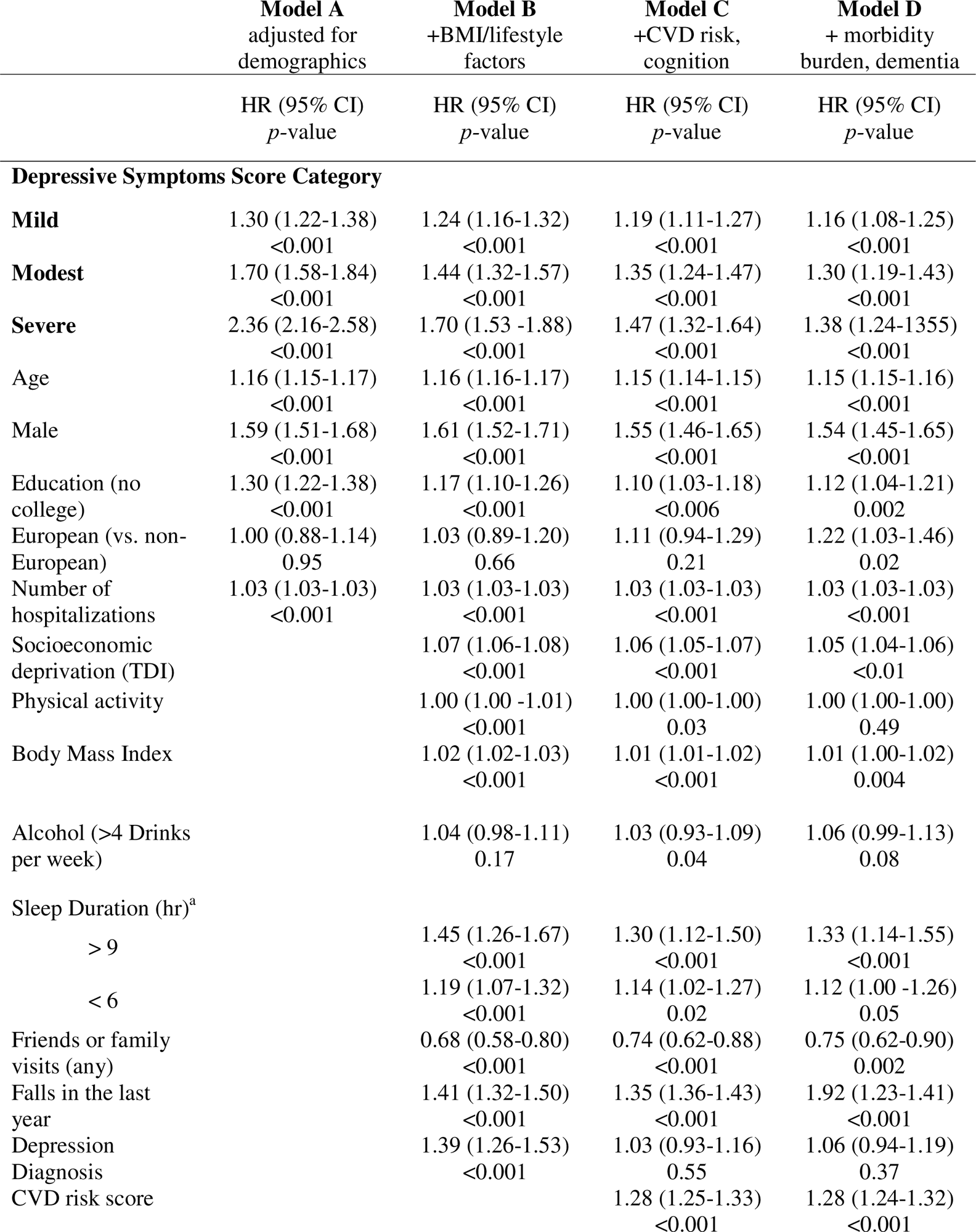
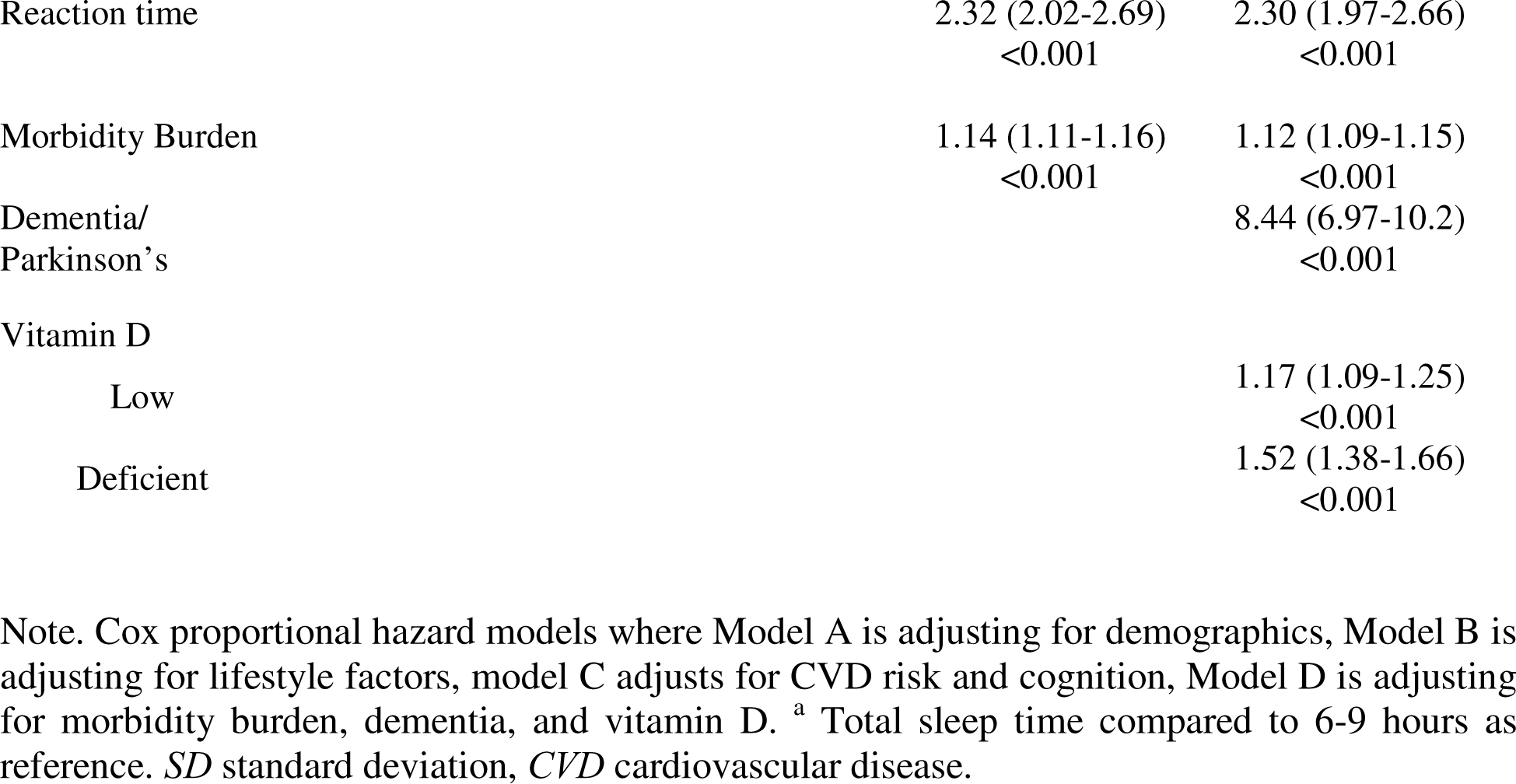
Full Cox Proportional Hazard Models.

**Supplemental Table 2.** Individual depression symptoms burden score components [daily vs. none] and risk for delirium.

|  | Model A<br>HR (95% CI)<br>*p* value | Model B<br>HR (95% CI)<br>*p* value | Model C<br>HR (95% CI)<br>*p* value | Model D<br>HR (95% CI)<br>*p* value |
| --- | --- | --- | --- | --- |
| Depressed mood | 2.17 (1.84-2.55)<br><0.001 | 1.37 (1.15-1.63)<br><0.001 | 1.25 (1.05-1.50)<br>0.01 | 1.15 (0.94-1.39)<br>0.17 |
| Unenthusiasm/<br>Disinterest | 1.88 (1.61-2.21)<br><0.001 | 1.15 (0.97-1.37)<br>0.10 | 1.08 (0.91-1.29)<br>0.38 | 1.06 (0.88-1.28)<br>0.56 |
| Tenseness/Restlessness | 2.25 (1.92-2.63)<br><0.001 | 1.52 (1.29-1.80)<br><0.001 | 1.42 (1.20-1.68)<br><0.001 | 1.34 (1.12-1.62)<br>0.002 |
| Tiredness/Lethargy | 2.48 (2.26-2.72)<br><0.001 | 1.64 (1.49-1.82)<br><0.001 | 1.45 (1.35-1.61)<br><0.001 | 1.41 (1.26-1.57)<br><0.001 |

Note. Individual components of Depression symptom burden score and hazard ratios for incident delirium comparing those who answered none vs daily for “frequency of ” in the past two weeks. Model A is a core model adjusting for demographics (age, sex, education, ethnic background, and number of hospitalizations. Model B includes Model A with Townsend deprivation index, physical activity, alcohol consumption, body mass index, sleep duration, frequency of friend and family visits, falls in the past year, and self-reported depression/anxiety or ICD depression diagnosis. Model C includes Model B with cardiovascular risk score, morbidity burden score, and reaction time. Model D includes Model C with dementia Parkinson’s diagnosis and vitamin D status \*[ Frequency in the past two weeks.]

**Supplemental Table 3.** Depressive symptoms burden score trajectory and risk for incident delirium within the Mild depressive symptoms burden category.

**Depressive Symptoms Burden Score trajectory\* (n = 162 with delirium)**
|  |  |  |  |
| --- | --- | --- | --- |
| No change/improved (0) | 28494 (75%) | Ref | Ref |
| Worsening (≥1) | 9650 (25%) | 1.45 (1.04-2.02) | 0.03 |

Note. Cox proportional hazard models comparing recent depressive symptoms score groups against reference group none, for all delirium cases using Model A, the core model adjusting for demographics (age, sex, education, ethnic background, and number of hospitalizations. Two-sided p value for HR in comparison with the reference category, without adjustment for multiple comparisons. \*Comparisons within the Mild group (0-2). Abbreviations: *HR* hazard ratio, *CI* confidence intervals.

## Supplemental Figures

**Supplemental Figure 1.**
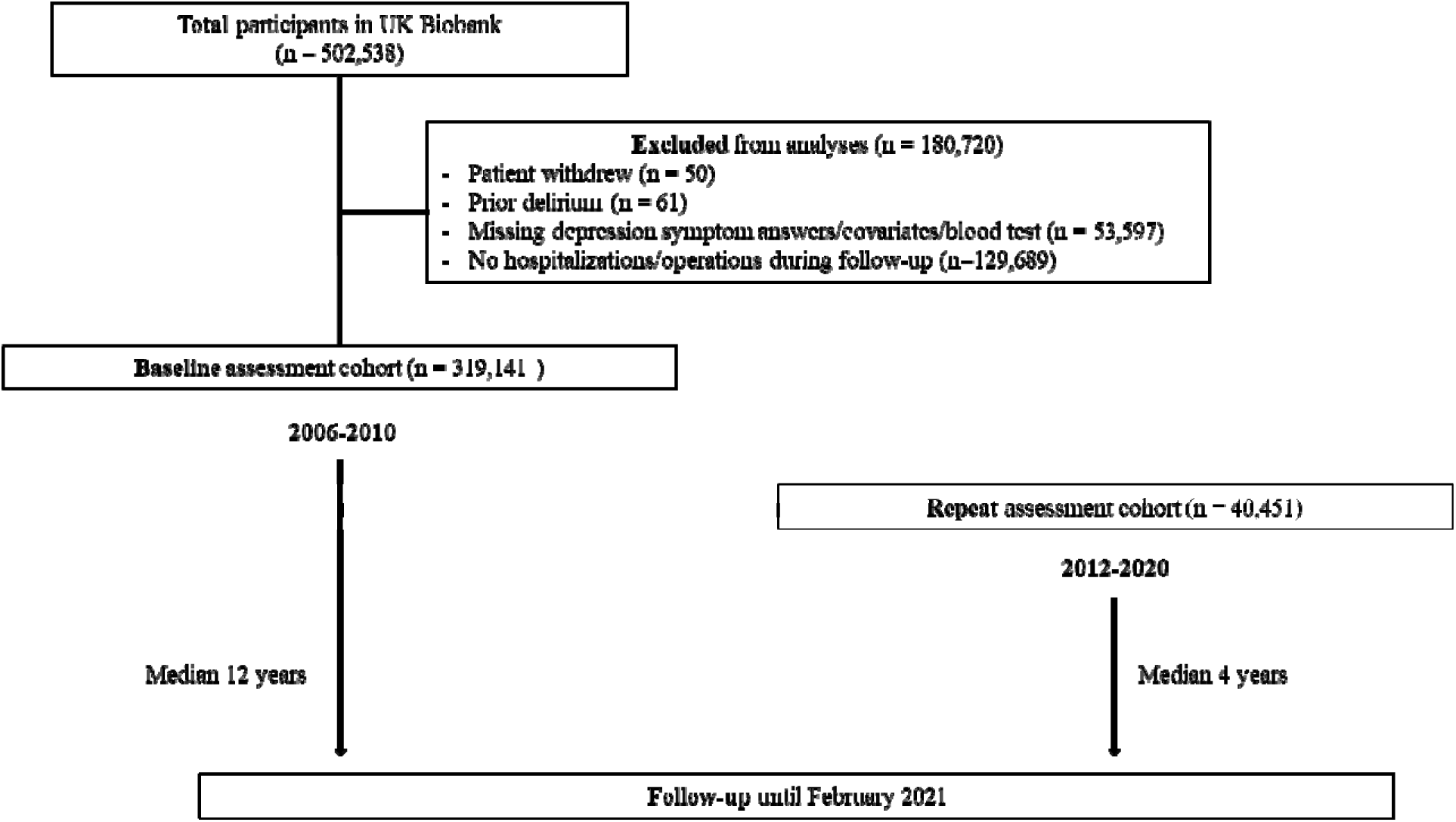
Flowchart of participant selection. How participants with incident delirium were selected in the study. From the UK biobank.

**Supplemental Figure 2.**
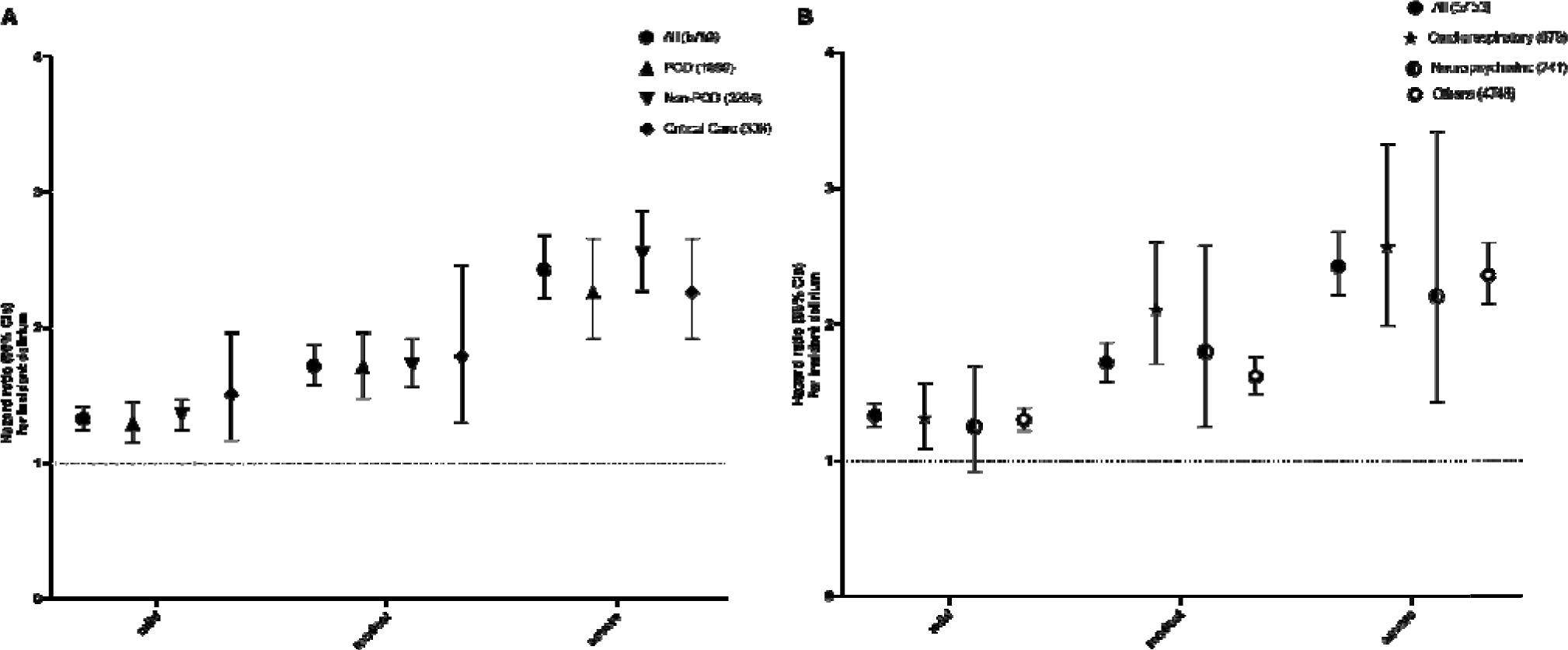
Cox proportional hazard models comparing the risk for incident delirium for each category of depression symptom burden score in each hospitalization setting (A), and admitting specialist/team (B). *POD* Post-operative delirium.

**Supplemental Figure 3.**
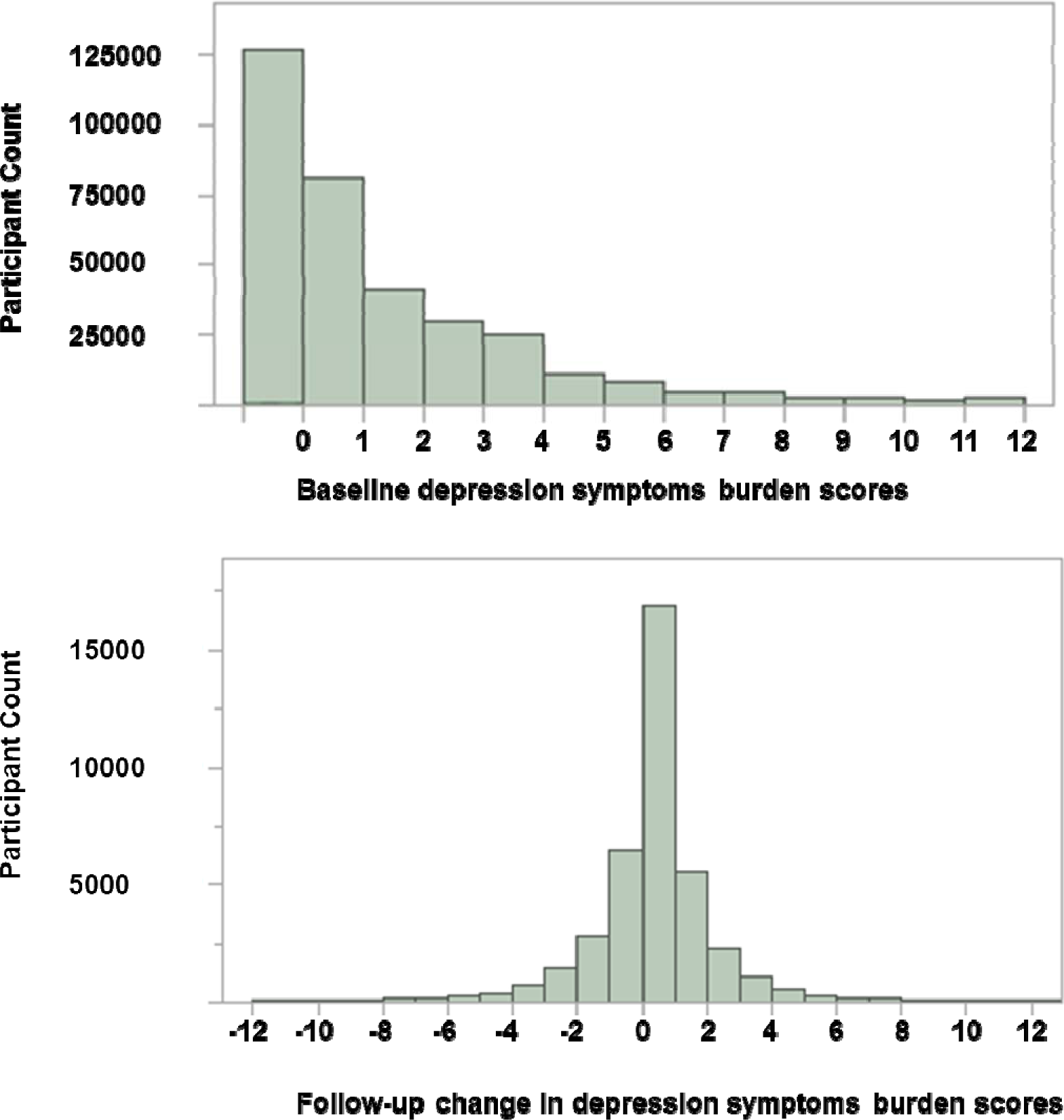
The number of participants for baseline depression symptoms burden score (0-12). Follow-up depression symptom burden score changes from baseline.

## References

1. Ryan DJ, O’Regan NA, Caoimh RÓ, et al. Delirium in an adult acute hospital population: predictors, prevalence and detection. BMJ Open. 2013;3(1). doi:10.1136/bmjopen-2012-001772

2. Siddiqi N, House AO, Holmes JD. Occurrence and outcome of delirium in medical in-patients: a systematic literature review. Age Ageing. 2006;35(4):350–364. doi:10.1093/ageing/afl005

3. Bickel H, Gradinger R, Kochs E, Förstl H. High Risk of Cognitive and Functional Decline after Postoperative Delirium. Dementia and Geriatric Cognitive Disorders. 2008;26(1):26–31. doi:10.1159/000140804

4. Ulsa MC, Xi Z, Li P, et al. Association of Poor Sleep Burden in Middle Age and Older Adults With Risk for Delirium During Hospitalization. J Gerontol A Biol Sci Med Sci. 2022;77(3):507–516. doi:10.1093/gerona/glab272

5. Leentjens AFG, Schieveld JNM, Leonard M, Lousberg R, Verhey FRJ, Meagher DJ. A comparison of the phenomenology of pediatric, adult, and geriatric delirium. Journal of Psychosomatic Research. 2008;64(2):219–223. doi:10.1016/j.jpsychores.2007.11.003

6. O’Sullivan R, Inouye SK, Meagher D. Delirium and depression: inter-relationship and clinical overlap in elderly people. Lancet Psychiatry. 2014;1(4):303–311. doi:10.1016/S2215-0366(14)70281-0

7. Givens JL, Jones RN, Inouye SK. The overlap syndrome of depression and delirium in older hospitalized patients. J Am Geriatr Soc. 2009;57(8):1347–1353. doi:10.1111/j.1532-5415.2009.02342.x

8. Cepoiu M, McCusker J, Cole MG, Sewitch M, Ciampi A. Recognition of depression in older medical inpatients. J Gen Intern Med. 2007;22(5):559–564. doi:10.1007/s11606-006-0085-0

9. Srifuengfung M, Abraham J, Avidan MS, Lenze EJ. Perioperative Anxiety and Depression in Older Adults: Epidemiology and Treatment. The American Journal of Geriatric Psychiatry. Published online July 8, 2023. doi:10.1016/j.jagp.2023.07.002

10. Smith PJ, Attix DK, Weldon BC, Greene NH, Monk TG. Executive function and depression as independent risk factors for postoperative delirium. In: The Journal of the American Society of Anesthesiologists. Vol 110. The American Society of Anesthesiologists; 2009:781–787.

11. Mychajliw C, Herrmann ML, Suenkel U, et al. Impaired Executive Function and Depression as Independent Risk Factors for Reported Delirium Symptoms: An Observational Cohort Study Over 8 Years. Frontiers in aging neuroscience. 2021;13:246.

12. Jankowski CJ, Trenerry MR, Cook DJ, et al. Cognitive and functional predictors and sequelae of postoperative delirium in elderly patients undergoing elective joint arthroplasty. Anesth Analg. 2011;112(5):1186–1193. doi:10.1213/ANE.0b013e318211501b

13. Detroyer E, Dobbels F, Verfaillie E, Meyfroidt G, Sergeant P, Milisen K. Is Preoperative Anxiety and Depression Associated with Onset of Delirium After Cardiac Surgery in Older Patients? A Prospective Cohort Study. Journal of the American Geriatrics Society. 2008;56(12):2278–2284. 10.1111/j.1532-5415.2008.02013.x

14. Fong TG, Davis D, Growdon ME, Albuquerque A, Inouye SK. The interface between delirium and dementia in elderly adults. Lancet Neurol. 2015;14(8):823–832. doi:10.1016/S1474-4422(15)00101-5

15. Chen H, Mo L, Hu H, Ou Y, Luo J. Risk factors of postoperative delirium after cardiac surgery: a meta-analysis. Journal of Cardiothoracic Surgery. 2021;16(1):113. doi:10.1186/s13019-021-01496-w

16. Enache D, Winblad B, Aarsland D. Depression in dementia: epidemiology, mechanisms, and treatment. Curr Opin Psychiatry. 2011;24(6):461–472. doi:10.1097/YCO.0b013e32834bb9d4

17. Sudlow C, Gallacher J, Allen N, et al. UK Biobank: An Open Access Resource for Identifying the Causes of a Wide Range of Complex Diseases of Middle and Old Age. PLoS Med. 2015;12(3):e1001779. doi:10.1371/journal.pmed.1001779

18. Allen N, Sudlow C, Downey P, et al. UK Biobank: Current status and what it means for epidemiology. Health Policy and Technology. 2012;1(3):123–126. doi:10.1016/j.hlpt.2012.07.003

19. Bowman K, Jones L, Pilling LC, et al. Vitamin D levels and risk of delirium: A mendelian randomization study in the UK Biobank. Neurology. 2019;92(12):e1387–e1394. doi:10.1212/WNL.0000000000007136

20. Pilling LC, Jones LC, Masoli JAH, et al. Low Vitamin D Levels and Risk of Incident Delirium in 351,000 Older UK Biobank Participants. Journal of the American Geriatrics Society. n/a(n/a). doi:10.1111/jgs.16853

21. Gao L, Gaba A, Li P, et al. Heart rate response and recovery during exercise predict future delirium risk-A prospective cohort study in middle- to older-aged adults. J Sport Health Sci. Published online December 13, 2021:S2095–2546(21)00140-X. doi:10.1016/j.jshs.2021.12.002

22. Gao L, Gaba A, Cui L, et al. Resting Heartbeat Complexity Predicts All-Cause and Cardiorespiratory Mortality in Middle- to Older-Aged Adults From the UK Biobank. J Am Heart Assoc. 2021;10(3):e018483. doi:10.1161/JAHA.120.018483

23. Whitlock EL, Vannucci A, Avidan MS. POSTOPERATIVE DELIRIUM. Minerva Anestesiol. 2011;77(4):448–456.

24. Sabia S, Fayosse A, Dumurgier J, et al. Association of sleep duration in middle and old age with incidence of dementia. Nature Communications. 2021;12(1):2289. doi:10.1038/s41467-021-22354-2

25. Gao L, Li P, Gaykova N, et al. Circadian Rest-Activity Rhythms, Delirium Risk, and Progression to Dementia. Ann Neurol. Published online February 20, 2023. doi:10.1002/ana.26617

26. Gao L, Zheng X, Baker SN, et al. Associations of rest-activity rhythm disturbances with stroke risk and post-stroke adverse outcomes. Published online May 16, 2023:2023.05.14.23289966. doi:10.1101/2023.05.14.23289966

27. Fawns-Ritchie C, Deary IJ. Reliability and validity of the UK Biobank cognitive tests. PLoS One. 2020;15(4). doi:10.1371/journal.pone.0231627

28. McAvay GJ, Van Ness PH, Bogardus Jr ST, et al. Depressive Symptoms and the Risk of Incident Delirium in Older Hospitalized Adults. Journal of the American Geriatrics Society. 2007;55(5):684–691. doi:10.1111/j.1532-5415.2007.01150.x

29. Greene NH, Attix DK, Weldon BC, Smith PJ, McDonagh DL, Monk TG. Measures of executive function and depression identify patients at risk for postoperative delirium. In: The Journal of the American Society of Anesthesiologists. Vol 110. The American Society of Anesthesiologists; 2009:788–795.

30. Smith PJ, Attix DK, Weldon BC, Monk TG. Depressive symptoms and risk of postoperative delirium. The American Journal of Geriatric Psychiatry. 2016;24(3):232–238.

31. American Geriatrics Society Expert Panel on Postoperative Delirium in Older Adults. Postoperative delirium in older adults: best practice statement from the American Geriatrics Society. J Am Coll Surg. 2015;220(2):136–148.e1. doi:10.1016/j.jamcollsurg.2014.10.019

32. Maldonado JR. Pathoetiological Model of Delirium: a Comprehensive Understanding of the Neurobiology of Delirium and an Evidence-Based Approach to Prevention and Treatment. Critical Care Clinics. 2008;24(4):789–856. doi:10.1016/j.ccc.2008.06.004

33. Jansson M, Gatz M, Berg S, et al. Association between depressed mood in the elderly and a5-HTR2A gene variant. Am J Med Genet. 2003;120B(1):79–84. doi:10.1002/ajmg.b.20016

34. Blazer DG, Burchett BB, Fillenbaum GG. APOE LJ4 and Low Cholesterol as Risks for Depression in a Biracial Elderly Community Sample. The American Journal of Geriatric Psychiatry. 2002;10(5):515–520. doi:10.1097/00019442-200209000-00004

35. O’Keeffe ST, Devlin JG. Delirium and the Dexamethasone Suppression Test in the Elderly. Neuropsychobiology. 1994;30(4):153–156. doi:10.1159/000119154

36. Kudoh A, Takase H, Katagai H, Takazawa T. Postoperative Interleukin-6 and Cortisol Concentrations in Elderly Patients with Postoperative Confusion. Neuroimmunomodulation. 2005;12(1):60–66. doi:10.1159/000082365

37. MacLullich AMJ, Ferguson KJ, Miller T, De Rooij SEJA, Cunningham C. Unravelling the pathophysiology of delirium: A focus on the role of aberrant stress responses. Journal of Psychosomatic Research. 2008;65(3):229–238. doi:10.1016/j.jpsychores.2008.05.019

38. Adamis D, Meagher D. Insulin-Like Growth Factor I and the Pathogenesis of Delirium: A Review of Current Evidence. Journal of Aging Research. 2011;2011:1–11. doi:10.4061/2011/951403

39. Alvaro PK, Roberts RM, Harris JK. A Systematic Review Assessing Bidirectionality between Sleep Disturbances, Anxiety, and Depression. Sleep. 2013;36(7):1059–1068. doi:10.5665/sleep.2810

40. Lim ASP, Saper CB. Sleep, circadian rhythms, and dementia. Ann Neurol. 2011;70(5):677–679. doi:10.1002/ana.22637

41. Fitzgerald JM, Adamis D, Trzepacz PT, et al. Delirium: A disturbance of circadian integrity? Medical Hypotheses. 2013;81(4):568–576. doi:10.1016/j.mehy.2013.06.032

42. Walker WH, Walton JC, DeVries AC, Nelson RJ. Circadian rhythm disruption and mental health. Translational psychiatry. 2020;10(1):1–13.

43. Daou M, Telias I, Younes M, Brochard L, Wilcox ME. Abnormal Sleep, Circadian Rhythm Disruption, and Delirium in the ICU: Are They Related? Front Neurol. 2020;11:549908. doi:10.3389/fneur.2020.549908

44. Prevention C for DC and. The state of aging and health in America 2013. In: CDC; 2018.

45. Selbæk G, Neerland BE. Cognitive decline and dementia: does delirium matter? The Lancet Healthy Longevity. 2022;3(4):e217–e218. doi:10.1016/S2666-7568(22)00056-3

46. Batty GD, Gale CR, Kivimäki M, Deary IJ, Bell S. Comparison of risk factor associations in UK Biobank against representative, general population based studies with conventional response rates: prospective cohort study and individual participant meta-analysis. BMJ. 2020;368:m131. doi:10.1136/bmj.m131

47. Givens JL, Jones RN, Inouye SK. The overlap syndrome of depression and delirium in older hospitalized patients. J Am Geriatr Soc. 2009;57(8):1347–1353. doi:10.1111/j.1532-5415.2009.02342.x

48. Lopez MN, Quan NM, Carvajal PM. A psychometric study of the Geriatric Depression Scale. European Journal of Psychological Assessment. Published online 2010.

49. Givens JL, Sanft TB, Marcantonio ER. Functional recovery after hip fracture: the combined effects of depressive symptoms, cognitive impairment, and delirium. Journal of the American Geriatrics Society. 2008;56(6):1075–1079.

50. Bowman K, Jones L, Pilling LC, et al. Vitamin D levels and risk of delirium: A mendelian randomization study in the UK Biobank. Neurology. 2019;92(12):e1387–e1394. doi:10.1212/WNL.0000000000007136

51. Sepulveda E, Franco JG, Trzepacz PT, et al. Delirium diagnosis defined by cluster analysis of symptoms versus diagnosis by DSM and ICD criteria: diagnostic accuracy study. BMC Psychiatry. 2016;16(1):167. doi:10.1186/s12888-016-0878-6

52. Casey P, Cross W, Mart MWS, Baldwin C, Riddell K, Dārziņš P. Hospital discharge data under-reports delirium occurrence: results from a point prevalence survey of delirium in a major Australian health service. Intern Med J. 2019;49(3):338–344. doi:10.1111/imj.14066

